# Association of the Covid-19 lockdown with smoking, drinking, and attempts to quit in England: an analysis of 2019-2020 data

**DOI:** 10.1101/2020.05.25.20112656

**Authors:** Sarah E. Jackson, Claire Garnett, Lion Shahab, Melissa Oldham, Jamie Brown

## Abstract

**Aim:** To examine changes in smoking, drinking, and quitting/reduction behaviour following the Covid-19 lockdown in England.

**Design/setting:** Monthly cross-sectional surveys representative of the adult population in England, aggregated before (April 2019 through February 2020) versus after (April 2020) the lockdown.

**Participants:** 20,558 adults (≥16y).

**Measurements:** The independent variable was the timing of the Covid-19 lockdown in England (before vs. after March 2020). Dependent variables were: prevalence of smoking and high-risk drinking; past-year cessation and quit attempts (among past-year smokers); past-year attempts to reduce alcohol consumption (among high-risk drinkers); and use of evidence-based (e.g., prescription medication/face-to-face behavioural support) and remote support (telephone support/websites/apps) for smoking cessation and alcohol reduction (among smokers/high-risk drinkers who made a quit/reduction attempt). Covariates included age, sex, social grade, region, and level of nicotine and alcohol dependence (as relevant).

**Findings:** The Covid-19 lockdown was not associated with a significant change in smoking prevalence (17.0% (after) vs. 15.9% (before), C)R=1.09[95%CI 0.95-1.24]), but was associated with increases in the rate of quit attempts (39.6% vs. 29.1%, OR_adj_=1.56[1.23-1.98]) and cessation (8.8% vs. 4.1%, OR_adj_=2.63[1.69-4.09]) among past-year smokers. Among smokers who tried to quit, there was no significant change in use of evidence-based support (50.0% vs. 51.5%, OR_a_dj=1.10[0.72-1.68]) but use of remote support increased (10.9% vs. 2.7%, OR_a_dj=3.59[1.56-8.23]). Lockdown was associated with increases in the prevalence of high-risk drinking (38.3% vs. 25.1%, OR=1.85[1.67-2.06]) but also alcohol reduction attempts by high-risk drinkers (28.5% vs. 15.3%, ORadr2.16[1.77-2.64]). Among high-risk drinkers who made a reduction attempt, use of evidence-based support decreased (1.2% vs. 4.0%, OR_a_dj=0.23[0.05-0.97]) and there was no significant change in use of remote support (6.9% vs. 6.1%, OR_adj_=1.32[0.64-2.75]).

**Conclusions:** In England, prevalence of high-risk drinking but not smoking has increased since the Covid-19 lockdown. Smokers and high-risk drinkers are more likely than before lockdown to report trying to quit smoking or reduce their alcohol consumption, and rates of smoking cessation are higher. Smokers are no less likely than before lockdown to use cessation support, with increased uptake of remote support. However, use of evidence-based support for alcohol reduction by high-risk drinkers has decreased, with no compensatory increase in use of remote support.

## Introduction

In the context of the Covid-19 pandemic, substance use remains a public health priority (1,2). Tobacco smoking and high-risk alcohol consumption are among the leading causes of disease and premature death worldwide (3,4). In England, approximately 14% of adults smoke (5) and 21% exceed UK drinking guidelines (6). Quitting smoking and reducing alcohol consumption can reduce the risk of chronic diseases and increase healthy life expectancy (7,8). Understanding what is happening to smoking, drinking, and quitting during the Covid-19 pandemic is important for evaluating the wider public health consequences of the pandemic. It also has important implications for informing the provision and targeting of support for smoking cessation and alcohol reduction.

Most governments have responded to the Covid-19 pandemic by advising the public to stay at home and avoid unnecessary social contact (so called ‘lockdown’ measures), to protect themselves and healthcare systems, and to save lives. The UK Coronavirus Action Plan (9) was published on 3 March 2020, followed by government advice to practice social distancing on 16 March and behavioural restrictions enforceable by law on 23 March. Daily news coverage of the rate of hospitalisations and deaths attributable to Covid-19 has emphasised its risks to health.

The Covid-19 pandemic, its associated health risks, and its impact on social activity may influence smoking and drinking in a number of ways. It may drive down the prevalence of smoking and high-risk drinking by providing a “teachable moment” that increases the salience of smoking- and alcohol-associated health risks and prompts people to make healthy changes to their behaviour (10). The disruption to daily routines caused by social distancing and stay-at-home (‘lockdown’) restrictions may reduce or eliminate usual smoking or drinking cues, making it easier to change these behaviours (11). Social smokers and drinkers may be less inclined to engage in the behaviours at home. Stay-at-home recommendations may also encourage cessation among smokers unable (e.g. because of rules set by a landlord or other family members) or unwilling (e.g. because they have children in the household) to smoke in the home.

On the other hand, there are reasons the lockdown may instead increase the prevalence of smoking and alcohol use and make quit attempts/reductions less of a priority. People are experiencing higher than usual levels of stress related to social isolation, employment, finances, caring responsibilities, and concerns about catching or becoming ill from the virus (12). Stress is an important risk factor for the onset and maintenance of alcohol misuse (13,14). Many smokers mistakenly believe that smoking helps to relieve stress and report smoking as a means of coping with high levels of stress (15,16). For those who are motivated to stop smoking or reduce their alcohol consumption, a (real or perceived) lack of support could provide a barrier to behaviour change. Under usual circumstances in England, a wide range of pharmacological and behavioural support is available for smoking cessation and alcohol reduction. Accessing such support is likely to be more difficult under social distancing and lockdown restrictions. While remote support (e.g. telephone support, websites, smartphone apps) that can be accessed from home is widely available, uptake by smokers and high-risk drinkers is low (17). A recent analysis of UK downloads of a popular smoking cessation app showed no evidence of a large increase in downloads in the period leading up to lockdown (18). Thus, by reducing access to popular methods of support, the lockdown may drive down the rate and/or success of quit attempts by smokers and reduction attempts by high-risk drinkers unless people switch to remote options.

While there are numerous reasons why the Covid-19 lockdown may have a positive or negative impact on both smoking and drinking, there are also key differences between these behaviours which mean we cannot presume that the net impact of the lockdown will be of the same magnitude, or even in the same direction. While Public Health England was advising smokers to quit to reduce their risk of worse Covid-19 outcomes (19), off-licenses were included in the list of essential business allowed to remain open during lockdown. There have been campaigns encouraging smokers to ‘QuitForCOVID’ (20) but no similar campaigns targeting alcohol. Social drinking may also be more likely than social smoking to continue during periods of social distancing, with people meeting online for virtual get-togethers, quizzes, etc. However, a potential protective effect of smoking (or nicotine) on Covid-19 risk has been widely publicised (21,22) which could increase smoking in the absence of an effect on drinking. Little evidence has been published on how the pandemic has affected smoking or alcohol consumption, but what evidence does exist suggests there may have been greater reductions in smoking than drinking. A survey of “1,000 people by YouGov and Action on Smoking and Health suggested an estimated 300,000 smokers in the UK had quit during the pandemic, while a further 550,000 had made a quit attempt (23,24). Meanwhile, a survey of “2,000 people by Alcohol Change UK suggested changes in drinking had been mixed, with one in three drinking less than usual during lockdown but one in five drinking more (25). Those who do report drinking more tend to be heavier drinkers (25).

There is a need for representative population-based data on how smoking, drinking, and quitting behaviour are affected by the Covid-19 lockdown. This study aimed to examine the extent to which smoking, drinking, and quitting/reduction behaviours have changed following the outbreak of Covid-19 in England among a representative sample. Specifically, we addressed the following research questions:

1. Among adults in England, has the prevalence of smoking or high-risk drinking changed following the outbreak of Covid-19, and if so, to what extent?
2. Among past-year smokers, has there been a change in the prevalence of cessation following the outbreak of Covid-19, after adjusting for sociodemographic characteristics and nicotine dependence?
3. Among past-year smokers, has there been a change in the prevalence of quit attempts following the outbreak of Covid-19, after adjusting for sociodemographic characteristics?
4. Among past-year smokers attempting to quit, has there been a change in the prevalence in the use of cessation support following the outbreak of Covid-19, after adjusting for sociodemographic characteristics and nicotine dependence?
5. Among high-risk drinkers, has there been a change in the prevalence of alcohol reduction attempts following the outbreak of Covid-19, after adjusting for sociodemographic characteristics?
6. Among high-risk drinkers attempting to reduce their alcohol consumption, has there been a change in the prevalence in the use of support for alcohol reduction following the outbreak of Covid-19, after adjusting for sociodemographic characteristics and alcohol dependence?

## Method

### Design

Data were drawn from the ongoing Smoking and Alcohol Toolkit Studies, monthly cross-sectional surveys of a representative sample of adults (≥16 years) in England designed to provide insights into population-wide influences on smoking and drinking behaviour (26,27). The studies use a form of random location sampling to select a new sample of approximately 1,700 adults aged ≥16 years each month. Interviews are performed with one household member until quotas based on factors influencing the probability of being at home (e.g. gender, age, working status) are fulfilled. Comparisons with sales data and other national surveys show that they recruit a representative sample of the population in England with regards to key demographic variables, smoking prevalence, and cigarette consumption (26,28). Data are usually collected monthly through face-to-face computer assisted interviews. However, social distancing restrictions under the Covid-19 lockdown meant that no data were collected in March 2020 and data from April 2020 were collected via telephone. The telephone-based data collection relied on the same combination of random location and quota sampling, and weighting approach as the face-to-face interviews.

For the present study, we used data from respondents to the survey in the period from April 2019 (one year before the height of the Covid-19 outbreak in the England) through April 2020 (the most recent data available at the time of analysis). We analysed aggregated data collected before (April 2019 through February 2020) versus after the lockdown (April 2020).

## Measures

### Smoking status

Smoking status was assessed with the question: “Which of the following best applies to you? *(a)* I smoke cigarettes (including hand-rolled) every day; (*b*) I smoke cigarettes (including hand-rolled), but not every day; *(c)* I do not smoke cigarettes at all, but I do smoke tobacco of some kind (e.g., pipe, cigar or shisha); (*d)* I have stopped smoking completely in the last year; *(e)* I stopped smoking completely more than a year ago; (*f*) I have never been a smoker (i.e. smoked for a year or more).” Current smoking was coded 1 for those who reported smoking any type of tobacco (i.e. responses *a-c)* and 0 for those who reported being a former or never smoker (responses *d-f)*. Past-year smoking was coded 1 for those who reported current smoking or having stopped in the past year (responses *a-d)* and 0 for those who reported stopping more than a year ago or never smoking (responses *e-f)*.

#### Smoking cessation

Among past-year smokers, cessation was coded 1 for those who reported having stopped smoking completely in the last year (response *e* to the measure of smoking status described above) and 0 for those who reported being a current smoker (responses *a-c)*.

### Attempts to stop smoking

Among past-year smokers, attempts to stop smoking was assessed with the question: “How many serious attempts to stop smoking have you made in the last 12 months? By serious attempt I mean you decided that you would try to make sure you never smoked again. Please include any attempt that you are currently making and please include any successful attempt made within the last year.” Those who reported making at least one serious quit attempt in the past year were coded 1, else they were coded 0.

### Use of support for smoking cessation

Among past-year smokers who reported making at least one quit attempt in the past year, use of cessation support in the most recent quit attempt as assessed with the question: “Which, if any, of the following did you try to help you stop smoking during the most recent serious quit attempt?” We analysed two variables: use of evidence-based support and use of remote support. Use of evidence-based support was coded 1 for those who reported using any of face-to-face behavioural support, prescription medication (varenicline, bupropion, or nicotine replacement therapy), e-cigarettes, or nicotine replacement therapy obtained over the counter, and 0 for those who did not report using any of these. Use of remote support was coded 1 for those who report using telephone support, a website, or an app, and 0 for those who did not report using any of these.

### High-risk drinking

Participants completed the three consumption questions of the Alcohol Use Disorders Identification Test (AUDIT-C) (29), a screening tool developed by the World Health Organization. The AUDIT-C classifies people scoring ≥5 as high-risk drinkers, and has demonstrated responsiveness to change, validity, high internal consistency, and good test-retest reliability across gender, age, and cultures (30–35).

### Attempts to restrict alcohol consumption

Among high-risk drinkers, attempts to reduce alcohol consumption were assessed with the question: “How many serious attempts to cut down on your drinking alcohol have you made in the last 12 months? By serious attempt I mean you decided that you would try to make sure you reduced the amount you drank permanently. Please include any attempt that you are currently making and please include any successful attempt made within the last 12 months.” Those who reported making at least one serious reduction attempt in the past year were coded 1, else they were coded 0.

### Use of support for alcohol reduction

Among high-risk drinkers who reported making at least one alcohol reduction attempt in the past year, use of support in the most recent attempt as assessed with the question: “Which, if any, of the following did you try to help you cut down during the most recent serious attempt?” We analysed two variables: use of evidence-based support and use of remote support. Use of evidence-based support was coded 1 for those who reported using any of face-to-face behavioural support or prescription medication (e.g. acamprosate, disulfiram, nalmefene), and 0 for those who did not report using any of these. Use of remote support was coded 1 for those who reported using telephone support, a website, or an app, and 0 for those who did not report using any of these.

### Covariates

Sociodemographic characteristics included age, sex, social grade, and region in England. Age was categorised as 16-24, 25-34, 35-44, 45-54, 55-64, and ≥65 years. Social grade was categorised as ABC1 (which includes managerial, professional and intermediate occupations) vs. C2DE (which includes small employers and own-account workers, lower supervisory and technical occupations, and semi-routine and routine occupations, never workers and long-term unemployed). This occupational measure of social grade is a valid index of SES, widely used in research in UK populations, which is particularly relevant in the context of tobacco use (36) and alcohol consumption (37). Regions in England were categorised as London, South, Central, and North.

We also included measures of nicotine and alcohol dependence. Nicotine dependence was assessed with the Heaviness of Smoking Index (38), an index derived from the number of cigarettes smoked per day and time to the first cigarette of the day. Scores range from 0 (low dependence) to 6 (high dependence). Alcohol dependence was assessed with the (full, 10-item) AUDIT (29). Scores range from 0-40, with 0-7 indicating low-risk consumption, 8-19 indicating hazardous or harmful consumption, and ≥20 indicating risk of alcohol dependence (moderate-severe alcohol use disorder).

## Statistical analysis

The study protocol and analysis plan were pre-registered on Open Science Framework (https://osf.io/q62k3). Analyses were done in SPSS v.24. Data were weighted to match the English population profile on age, social grade, region, tenure, ethnicity, and working status within sex. The dimensions are derived monthly from a combination of the English 2011 census, Office for National Statistics mid-year estimates, and an annual random probability survey conducted for the National Readership Survey. Missing cases were excluded on a per-analysis basis.

We used descriptive statistics and logistic regression to estimate the prevalence and odds of (i) current smoking, (ii) cessation by smokers, (iii) quit attempts by smokers; (iv) use of cessation support by smokers who made a quit attempt, (v) high-risk drinking, (vi) attempts to reduce alcohol consumption by high-risk drinkers, and (vii) use of support by high-risk drinkers making a reduction attempt, in relation to the timing of the Covid-19 outbreak in England (before [referent] vs. after). Estimates of smoking and high-risk drinking prevalence are reported unadjusted (as they were weighted on important dimensions to match the population in England). Analyses of quit/reduction attempts are reported with and without adjustment for age, sex, social grade, and region (to take account of small differences in the make-up of the subgroups being analysed). Analyses of smoking cessation and use of support are reported with and without adjustment for sociodemographic characteristics and level of dependence (because more dependent smokers tend to be less likely to quit and more dependent smokers/drinkers tend to be more likely to use support).

Because we had just one wave of data collected (and thus a relatively small sample) after the Covid-19 lockdown (April 2020), we did not explore moderating effects via interactions between the timing of the lockdown and sociodemographic characteristics or level of dependence. However, in Supplementary File 1 we report the prevalence of smoking (Table 1) and drinking (Table 2) outcomes by age, sex, social grade, region, and level of dependence. When more data have been collected after the start of the lockdown and statistical power is sufficient to detect significant interactions, this is something we will examine in more detail in a separate paper.

**Table 1.** Characteristics of the samples recruited before and after the Covid-19 lockdown

|  | Before lockdown<br>(April 2019-February 2020) | After lockdown<br>(April 2020) | *p* |
| --- | --- | --- | --- |
| *N* | 18884 | 1674 | - |
| Age in years, % (*n*) |  |  |  |
| 16-24 | 13.4 (2538) | 10.9 (183) | 0.134 |
| 25-34 | 16.9 (3187) | 17.2 (288) | - |
| 35-44 | 15.7 (2964) | 16.0 (268) | - |
| 45-54 | 17.0 (3208) | 17.0 (285) | - |
| 55-64 | 14.5 (2746) | 15.2 (255) | - |
| ≥65 | 22.5 (4241) | 23.3 (389) | - |
| Missing | 0 (0) | 0.4 (6) | - |
| Sex, % (*n*) |  |  |  |
| Male | 49.1 (9273) | 49.1 (822) | 0.999 |
| Female | 50.9 (9611) | 50.9 (852) | - |
| Social grade, % (*n*) |  |  |  |
| ABC1 | 55.4 (10457) | 53.7 (898) | 0.705 |
| C2DE | 44.6 (8427) | 44.1 (738) | - |
| Missing | 0 (0) | 2.3 (38) |  |
| Region in England, % (*n*) |  |  |  |
| London | 15.7 (2971) | 15.8 (264) | 0.994 |
| South | 26.4 (4983) | 26.5 (444) | - |
| Central | 30.1 (5692) | 30.3 (507) | - |
| North | 27.7 (5238) | 27.4 (459) | - |

Note: All data are weighted to match the adult population in England on age, social grade, region, tenure, ethnicity, and working status within sex.

**Table 2.**
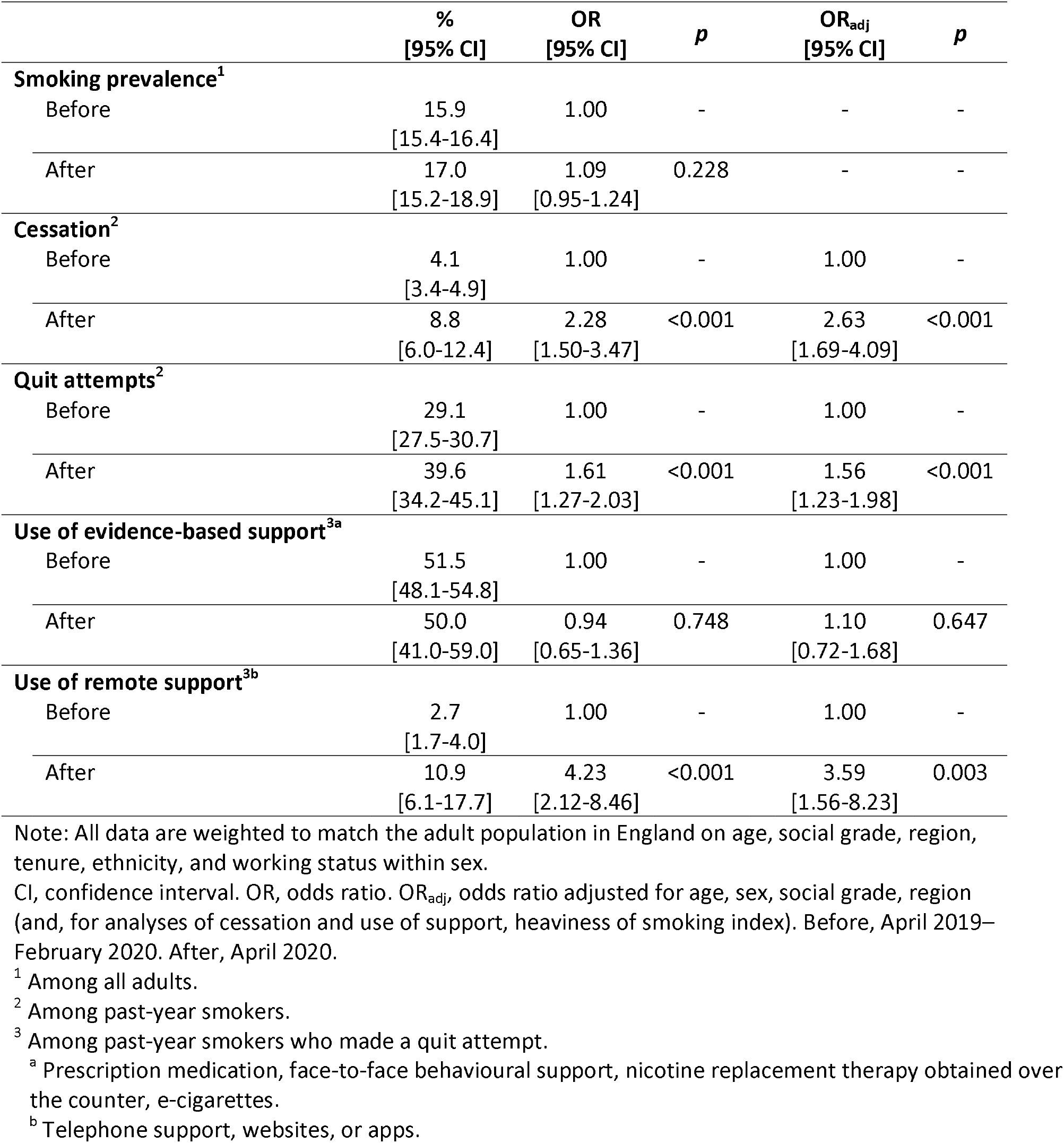
Association of the timing of the Covid-19 lockdown with smoking and quitting behaviour in England

|  | %<br>[95% CI] | OR<br>[95% CI] | *p* | OR<sub>adj</sub><br>[95% CI] | *p* |
| --- | --- | --- | --- | --- | --- |
| **Smoking prevalence<sup>1</sup>** |  |  |  |  |  |
| Before | 15.9<br>[15.4-16.4] | 1.00 | - | - | - |
| After | 17.0<br>[15.2-18.9] | 1.09<br>[0.95-1.24] | 0.228 | - | - |
| **Cessation<sup>2</sup>** |  |  |  |  |  |
| Before | 4.1<br>[3.4-4.9] | 1.00 | - | 1.00 | - |
| After | 8.8<br>[6.0-12.4] | 2.28<br>[1.50-3.47] | <0.001 | 2.63<br>[1.69-4.09] | <0.001 |
| **Quit attempts<sup>2</sup>** |  |  |  |  |  |
| Before | 29.1<br>[27.5-30.7] | 1.00 | - | 1.00 | - |
| After | 39.6<br>[34.2-45.1] | 1.61<br>[1.27-2.03] | <0.001 | 1.56<br>[1.23-1.98] | <0.001 |
| **Use of evidence-based support<sup>3a</sup>** |  |  |  |  |  |
| Before | 51.5<br>[48.1-54.8] | 1.00 | - | 1.00 | - |
| After | 50.0<br>[41.0-59.0] | 0.94<br>[0.65-1.36] | 0.748 | 1.10<br>[0.72-1.68] | 0.647 |
| **Use of remote support<sup>3b</sup>** |  |  |  |  |  |
| Before | 2.7<br>[1.7-4.0] | 1.00 | - | 1.00 | - |
| After | 10.9<br>[6.1-17.7] | 4.23<br>[2.12-8.46] | <0.001 | 3.59<br>[1.56-8.23] | 0.003 |

Note: All data are weighted to match the adult population in England on age, social grade, region, tenure, ethnicity, and working status within sex.
CI, confidence interval. OR, odds ratio. OR<sub>adj</sub>, odds ratio adjusted for age, sex, social grade, region (and, for analyses of cessation and use of support, heaviness of smoking index). Before, April 2019–February 2020. After, April 2020.
<sup>1</sup> Among all adults.
<sup>2</sup> Among past-year smokers.
<sup>3</sup> Among past-year smokers who made a quit attempt.
<sup>a</sup> Prescription medication, face-to-face behavioural support, nicotine replacement therapy obtained over the counter, e-cigarettes.
<sup>b</sup> Telephone support, websites, or apps.

In order to evaluate the potential impact of the change in modality of data collection from face-to-face (before the lockdown) to telephone (after the lockdown started) on the representativeness of the sample, and thus comparability of data, we conducted a series of diagnostic analyses (Supplementary File 2). While these identified some differences in the unweighted sociodemographic profiles of the face-to-face and telephone samples, the weighting required to achieve a representative sample was similar across modalities, and expected associations between smoking, high-risk drinking, and sociodemographic characteristics were observed on unweighted data in the telephone sample. Moreover, previous studies that have compared face-to-face and telephone interviews have demonstrated a high degree of comparability (39,40). This suggests it is reasonable to compare data from before and after the lockdown, despite the change in data collection method.

## Results

A total of 18,884 adults aged ≥18 years participated in the Smoking Toolkit Study between April 2019 and February 2020 (mean [SD] 1,717 [35.3] per month) and 1,674 participated in April 2020. Sociodemographic characteristics of the two samples are shown in Table 1.

Table 2 shows the prevalence and odds of current smoking, cessation, quit attempts, and use of cessation support before and after the Covid-19 lockdown. Figure 1 shows the monthly prevalence of these outcomes across the study period. Among adults in England, there was no significant difference in smoking prevalence after compared with before the lockdown (17.0% vs. 15.9%). However, among past-year smokers, odds of quitting were 2.63 times higher and odds of attempting to quit were 1.56 times higher after compared with before the lockdown started, when adjusting for covariates. Among past-year smokers who attempted to quit, odds of using remote cessation support was 3.59 times higher after compared with before the lockdown, when adjusting for covariates, but odds of using evidence-based support did not differ significantly.

**Figure 1.**
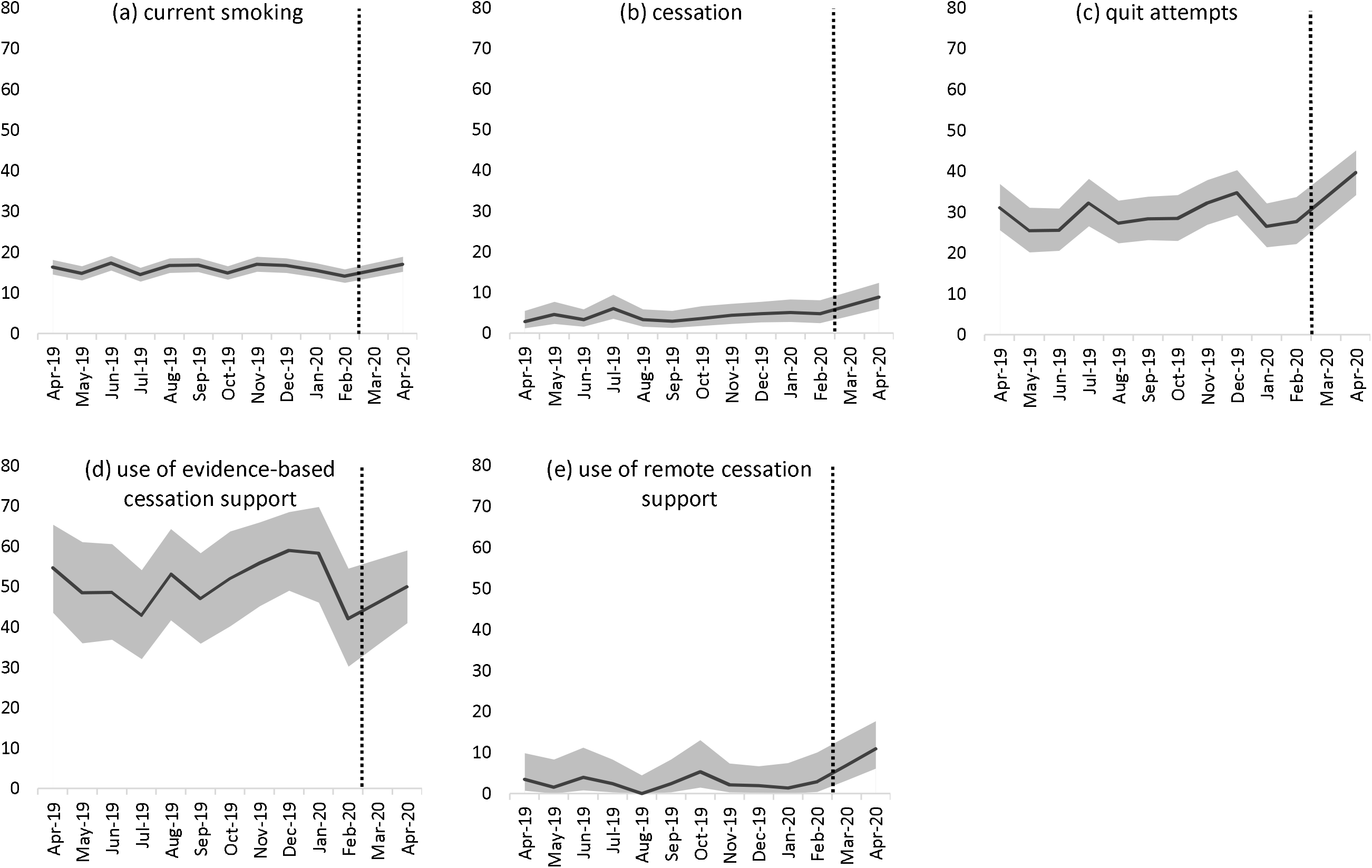
Prevalence of (a) current smoking among all adults; (b) cessation and (c) quit attempts by past-year smokers; and (d) use of evidence-based cessation support and (e) use of remote cessation support by past-year smokers who made a quit attempt in England, April 2019 through April 2020. The dotted line indicates the timing of the Covid-19 lockdown in England. Data for March 2020 were imputed as the average of February and April 2020 on the basis of presumed linear change.

Table 3 shows the prevalence and odds of high-risk drinking, alcohol reduction attempts, and use of support for alcohol reduction before and after the Covid-19 lockdown. Figure 2 shows the monthly prevalence of these outcomes across the study period. Among adults in England, the prevalence of high-risk drinking was significantly higher after compared with before the lockdown (38.3% vs. 25.1%; OR 1.85). Among high-risk drinkers, odds of making a serious attempt to reduce alcohol consumption were 2.16 times higher after than before the lockdown, when adjusting for covariates. Among high-risk drinkers who made a reduction attempt, odds of using evidence-based support was 0.23 times lower after compared with before the lockdown, when adjusting for covariates, but odds of using remote support did not differ significantly.

**Table 3.** Association of the timing of the Covid-19 lockdown with high-risk drinking and alcohol reduction attempts in England

|  | %<br>[95% CI] | OR<br>[95% CI] | *p* | OR<sub>adj</sub><br>[95% CI] | *p* |
| --- | --- | --- | --- | --- | --- |
| **High-risk drinking prevalence<sup>1</sup>** |  |  |  |  |  |
| Before | 25.1<br>[24.4-25.7] | 1.00 | - | - | - |
| After | 38.3<br>[35.9-40.7] | 1.85<br>[1.67-2.06] | <0.001 | - | - |
| **Alcohol reduction attempts<sup>2</sup>** |  |  |  |  |  |
| Before | 15.3<br>[14.3-16.4] | 1.00 | - | 1.00 | - |
| After | 28.5<br>[25.0-32.3] | 2.21<br>[1.82-2.68] | <0.001 | 2.16<br>[1.77-2.64] | <0.001 |
| **Use of evidence-based support<sup>3a</sup>** |  |  |  |  |  |
| Before | 4.0<br>[2.7-5.7] | 1.00 | - | 1.00 | - |
| After | 1.2<br>[0.1-4.1] | 0.32<br>[0.08-1.26] | 0.103 | 0.23<br>[0.05-0.97] | 0.046 |
| **Use of remote support<sup>3b</sup>** |  |  |  |  |  |
| Before | 6.1<br>[4.5-8.2] | 1.00 | - | 1.00 | - |
| After | 6.9<br>[3.6-11.8] | 1.21<br>[0.63-2.32] | 0.573 | 1.32<br>[0.64-2.75] | 0.456 |

Note: All data are weighted to match the adult population in England on age, social grade, region, tenure, ethnicity, and working status within sex.
CI, confidence interval. OR, odds ratio. OR<sub>adj</sub>, odds ratio adjusted for age, sex, social grade, region (and, for analyses of use of support, full AUDIT score as an indicator of dependence). Before, April 2019–February 2020. After, April 2020.
<sup>1</sup> Among all adults.
<sup>2</sup> Among high-risk drinkers.
<sup>3</sup> Among high-risk drinkers who made a reduction attempt.
<sup>a</sup> Prescription medication or face-to-face behavioural support.
<sup>b</sup> Telephone support, websites, or apps.

**Figure 2.**
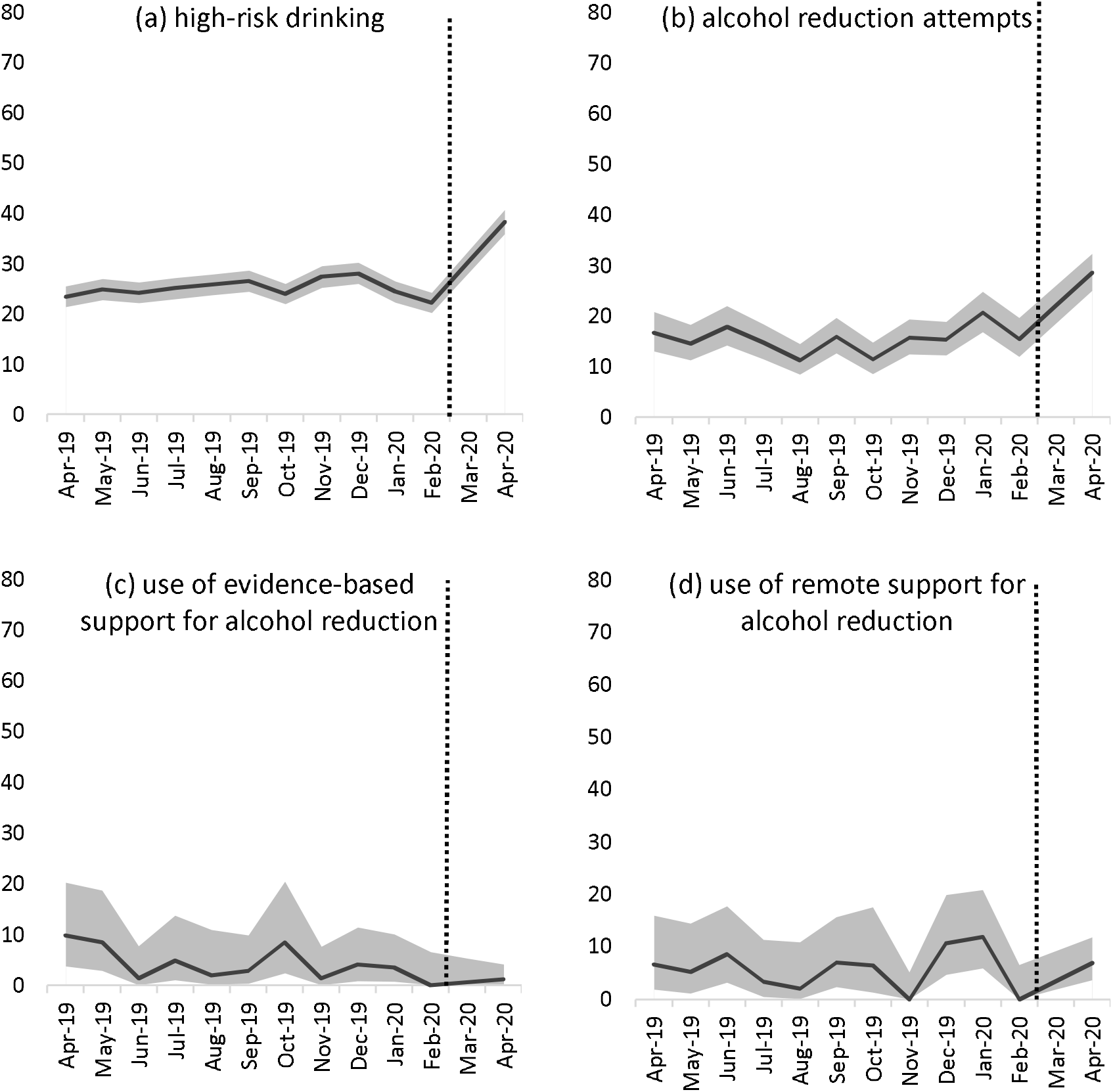
Prevalence of (a) high-risk drinking among all adults; (b) reduction attempts by high-risk drinkers; and (c) use of evidence-based support and (d) use of remote support for alcohol reduction by high-risk drinkers who made a reduction attempt in England, April 2019 through April 2020. The dotted line indicates the timing of the Covid-19 lockdown in England. Data for March 2020 were imputed as the average of February and April 2020 on the basis of presumed linear change.

## Discussion

Using a series of monthly surveys representative of adults in England, we examined changes in smoking, drinking, quitting and reduction attempts from before (April 2019-February 2020) to after (April 2020) the Covid-19 lockdown was implemented. Results showed the lockdown was not associated with a significant change in smoking prevalence, but was associated with increases in cessation and quit attempts by smokers. Among smokers who tried to quit, there was no significant change in use of evidence-based support but use of remote support increased. The lockdown was associated with increases in the prevalence of high-risk drinking but also with alcohol reduction attempts by high-risk drinkers. Among high-risk drinkers who made a reduction attempt, use of evidence-based support decreased and there was no significant change in use of remote support.

That smoking prevalence did not increase significantly in response to the Covid-19 lockdown is encouraging. Descriptive studies of patients hospitalised with Covid-19 have documented disproportionately low rates of current smoking compared with those observed in the general population (41-43), which has led to speculation that nicotine may be protective against adverse Covid-19 outcomes (44,45), and media coverage that smokers may be protected (21,22). Our results suggest that there has not been substantial uptake of or relapse to smoking despite this widespread coverage.

The observed increase in high-risk drinking, on the other hand, provides cause for concern – both in the context of Covid-19 risk and public health more broadly. Excessive alcohol consumption may increase the risk of Covid-19 directly, via adverse immune-related health effects (46), or indirectly, via reduced vigilance (47) around social distancing and adherence to other protective behaviours. An increase in high-risk drinking is also likely to put increased strain on health services which are already stretched to capacity under the lockdown. Our results add weight to calls for warnings around the risks of excessive drinking during isolation to be included in public health messaging related to the pandemic (2).

While smoking prevalence remained fairly stable and high-risk drinking increased, significant increases were observed in the proportion of smokers quitting and making quit attempts, and the proportion of high-risk drinkers attempting to reduce their alcohol consumption. Potential explanations for these changes include that the Covid-19 pandemic and lockdown provided a “teachable moment” that prompted healthy behaviour change, or changes in usual daily routines and social activities providing the opportunity to change smoking and drinking behaviours. While there are also reasons why the lockdown may suppress smoking cessation or alcohol reduction - for example, due to increased stress levels (12) - it appears that the net impact of the Covid-19 lockdown is one of increased effort to quit smoking and drink less (although the latter must be considered in the context of increased levels of high-risk drinking and may reflect attempts to return to usual levels of consumption).

Patterns in use of support differed by behaviour. The Covid-19 lockdown was associated with no change in use of evidence-based support for smoking cessation (prescription medication, face-to-face behavioural support, e-cigarettes, over-the-counter nicotine replacement therapy) but an increase in use of remote support (telephone support, websites, or apps). However, for alcohol reduction, the lockdown was associated with a reduction in use of evidence-based support (prescription medication, face-to-face behavioural support) and no change in use of remote support. It is possible that smokers were more able than high-risk drinkers to access evidence-based support in the form of e-cigarettes and nicotine replacement therapy, which are available online. However, there is no reason why high-risk drinkers should not have sought out remote support when traditional methods of support were less accessible. It is possible that the increase in use of remote support by smokers was attributable to campaigns on social media directing smokers to relevant websites and apps (e.g. 20); we are not aware of equivalent campaigns for alcohol (some people suggested a ‘Dry Covid’ could be beneficial (48), but this did not gain traction). Information campaigns on the range of support available for drinkers who wish to reduce their alcohol consumption could be useful in increasing awareness of remote options.

This study had several strengths, including the repeat cross-sectional design across the key time period, representative sample, and breadth of data collected on smoking, drinking, quitting, and alcohol reduction. However, there were also limitations. There was a change in the modality of data collection from face-to-face (before the lockdown) to telephone (after the lockdown started), which may be associated with the changes observed rather than the inferred association with the lockdown. However, we ran diagnostic analyses to compare the representativeness of the sample before and after the modality change, which suggested the comparisons were reasonable. While we identified some differences in the unweighted sociodemographic profiles of the face-to-face and telephone samples, the weighting required to achieve a representative sample was similar across modalities, and we observed expected associations between smoking, high-risk drinking, and sociodemographic characteristics on unweighted data in the telephone sample. A second limitation is that with only one wave of post-lockdown data collected to date, this study provides a simple assessment of changes in the prevalence of key indicators of smoking and alcohol use. The optimal design to evaluate the impact of the Covid-19 lockdown on these behaviours is an interrupted time series design, which models the effect of an intervention (in this case, the lockdown), taking account of long-term trends in the data. This will not be possible for at least a year. Given the importance of health behaviours for public health, and the need for up-to-date information in this unprecedented health and social landscape, we believed it was important to provide initial results now, and conduct a more sophisticated time series analysis when sufficient data points are available. A final limitation is that quitting and reduction activity was assessed in the context of the last 12 months. As a result, prevalence estimates of smoking cessation, quit attempts, alcohol reduction attempts, and use of support reflect activity over the past year and, in some cases, may have occurred before the lockdown. Also, our analysis does not account for seasonal differences in these behaviours. Caution should therefore be taken in extrapolating our results to provide estimates of the total number of smokers or high-risk drinkers who have tried to quit in response to the Covid-19 pandemic and lockdown. However, the timing of these outcomes should not affect estimates of the association of the lockdown with quitting/reduction behaviour because it affects the pre- and post-Covid-19 samples equally.

In conclusion, the prevalence of high-risk drinking in England has increased since the Covid-19 lockdown, but prevalence of smoking remains similar. Smokers and high-risk drinkers are more likely than before the lockdown to report trying to quit smoking or reduce their alcohol consumption, and rates of smoking cessation are higher. Smokers are no less likely than before the lockdown to use cessation support, with increased uptake of remote support (e.g. quitlines, websites, and apps). However, use by high-risk drinkers of evidence-based support for alcohol reduction has decreased, with no compensatory increase in use of remote support.

## Data Availability

Data are available from the corresponding author upon request.

## Declarations

### Ethics approval and consent to participate

Ethical approval for the Smoking Toolkit Study was granted originally by the UCL Ethics Committee (ID 0498/001). The data are not collected by UCL and are anonymised when received by UCL.

### Availability of data and materials

Data are available from the corresponding author upon request.

